# Artificial Intelligence-Derived Extracellular Volume Fraction for Diagnosis and Prognostication in Patients with Light-Chain Cardiac Amyloidosis

**DOI:** 10.1101/2024.03.20.24304642

**Authors:** In-Chang Hwang, Eun Ju Chun, Pan Ki Kim, Myeongju Kim, Jiesuck Park, Hong-Mi Choi, Yeonyee E. Yoon, Goo-Yeong Cho, Byoung Wook Choi

## Abstract

**Aims:** T1 mapping on cardiac magnetic resonance (CMR) imaging is useful for diagnosis and prognostication in patients with light-chain cardiac amyloidosis (AL-CA). We conducted this study to evaluate the performance of T1 mapping parameters for detection of cardiac amyloidosis (CA) in patients with left ventricular hypertrophy (LVH) and their prognostic values in patients with AL-CA, using a semi-automated deep learning algorithm.

**Methods and Results:** A total of 300 patients who underwent CMR for differential diagnosis of LVH were analyzed. CA was confirmed in 50 patients (39 with AL-CA and 11 with transthyretin amyloidosis), hypertrophic cardiomyopathy in 198, hypertensive heart disease in 47, and Fabry disease in 5. A semi-automated deep learning algorithm (Myomics-Q) was used for the analysis of the CMR images. The optimal cutoff extracellular volume fraction (ECV) for the differentiation of CA from other etiologies was 33.6% (diagnostic accuracy 85.6%). he artificial intelligence (AI)-derived ECV showed a significant prognostic value for a composite of cardiovascular death and heart failure hospitalization in patients with AL-CA (revised Mayo stage III or IV) (adjusted hazard ratio 4.247 for ECV ≥40%, 95% confidence interval 1.215–14.851, p-value=0.024). Incorporation of AI-derived ECV into the revised Mayo staging system resulted in better risk stratification (integrated discrimination index 27.9%, p=0.013; net reclassification index 13.8%, p=0.007).

**Conclusions:** AI-assisted T1 mapping on CMR imaging allows for improved diagnosis of CA from other etiologies of LVH. Furthermore, AI-derived ECV has significant prognostic value in patients with AL-CA, suggesting its clinical usefulness.

**Graphical Abstract:** 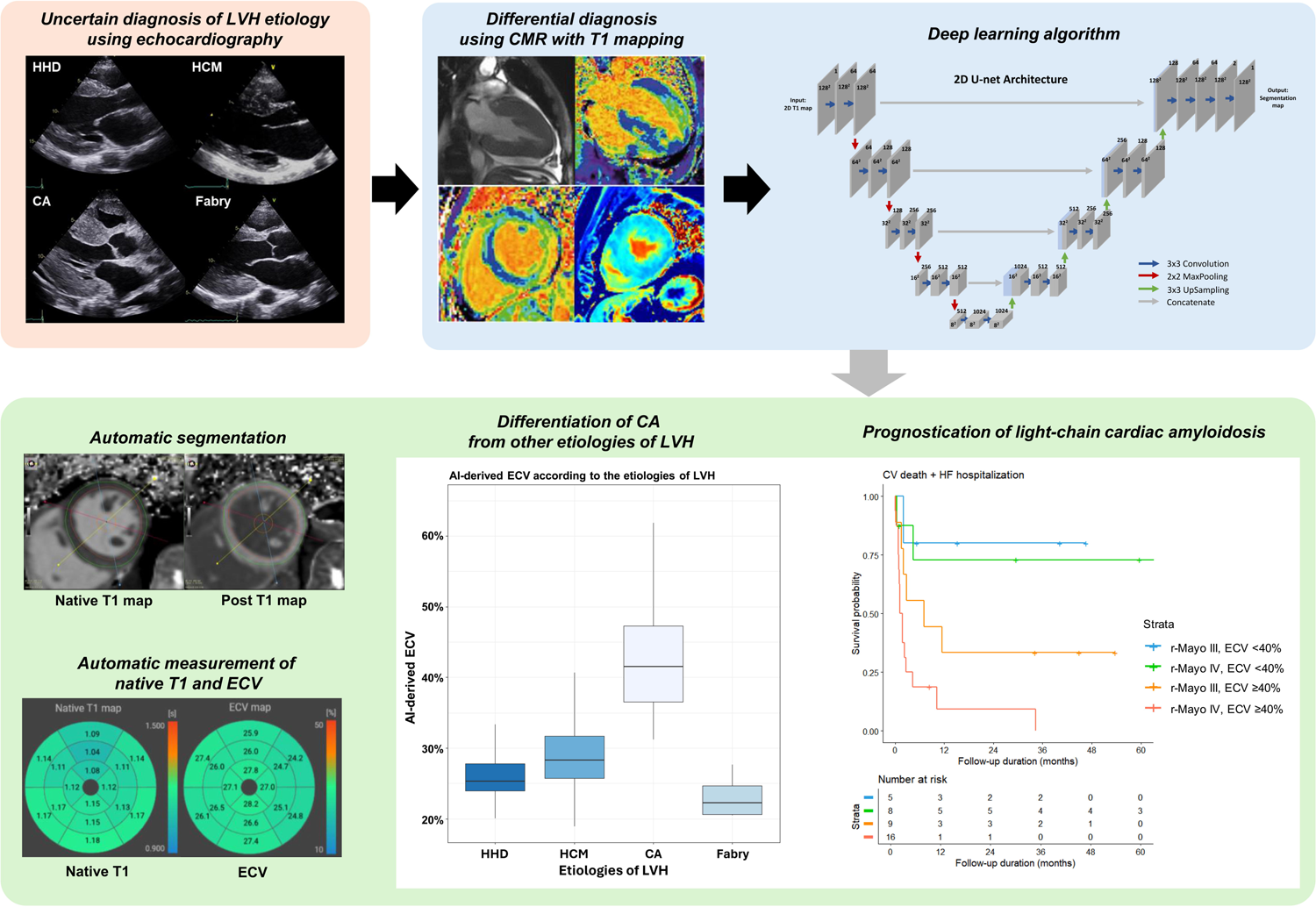

## Introduction

Differential diagnosis of left ventricular hypertrophy (LVH) is one of the most challenging issues in cardiovascular imaging.(1) The different etiologies of LVH have morphological similarities, but unique pathophysiology, treatment strategy, and prognosis.(2, 3) In particular, differential diagnosis of cardiac amyloidosis (CA) is critical for effective management: light-chain cardiac amyloidosis (AL-CA) is a hematologic malignancy that requires cytotoxic chemotherapy and/or stem cell transplantation, whereas transthyretin CA (TTR-CA) is a rare infiltrative cardiomyopathy that requires specific treatments such as TTR stabilization.(4)

While several combinations of clinical, electrocardiographic, and echocardiographic features have shown acceptable accuracy, cardiac magnetic resonance (CMR) remains the most important imaging modality for the differential diagnosis of LVH.(4) The presence and patterns of late gadolinium enhancement (LGE) have proven to be relevant markers for the detection of CA and hypertrophic cardiomyopathy (HCM) from hypertensive heart disease (HHD).(2, 5) In addition, novel parameters of myocardial fibrosis, including native T1 and extracellular volume fraction (ECV), have proven to be potentially useful for differential diagnosis and prognostication in patients with various etiologies of LVH, especially in those with CA.(6)

Despite the rapid progress of artificial intelligence (AI) in CMR, there has been no study that showed the usefulness of AI in the T1 mapping on CMR images of patients with LVH. Therefore, we aimed to assess the diagnostic performance of an AI-assisted semi-automated T1 mapping for the detection of CA in patients with LVH. In addition, we analyzed the prognostic value of the AI-derived ECV in patients with AL-CA in comparison with current clinical staging system.

## Methods

### Study design and cohort

This study was conducted in accordance with the principles outlined in the 2013 revised Declaration of Helsinki and approved by Seoul National University Bundang Hospital Institutional Review Board. The requirement for informed consent was waived owing to the retrospective nature of the study and the minimal expected risk to the patients.

We retrospectively identified 300 patients (47 with HHD, 198 with HCM, 50 with CA [39 with AL-CA, and 11 with TTR-CA], and 5 with Fabry disease [FD]) (**Figure 1**) who underwent CMR for differential diagnosis of the etiology, based on the detection of LVH on echocardiography. The diagnostic criteria for HHD, HCM, AL-CA, TTR-CA, and FD are described below.

**Figure 1.**
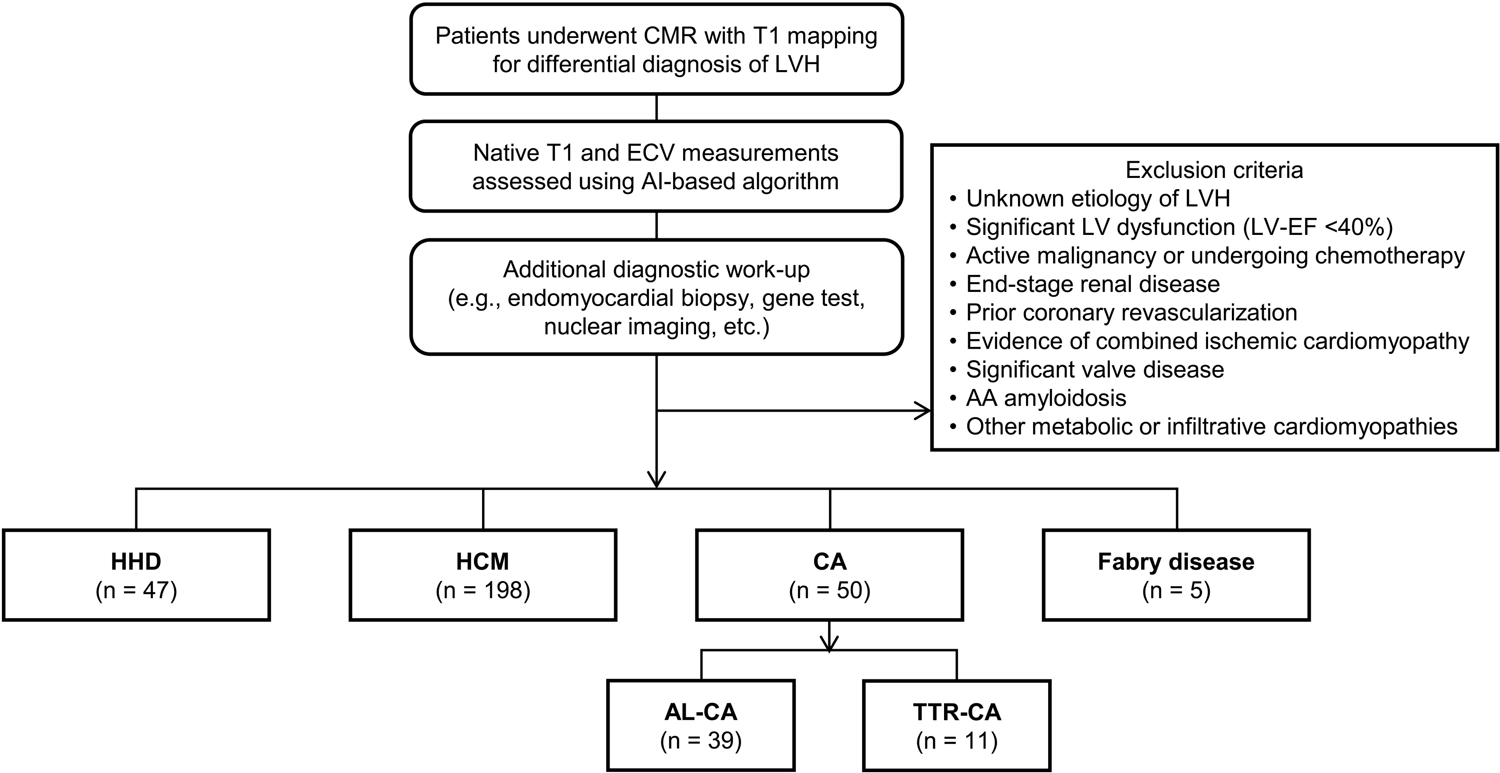
Flowchart of patient selection Abbreviations: CMR, cardiac magnetic resonance; LVH, left ventricular hypertrophy; ECV, extracellular volume fraction; HHD, hypertensive heart disease; HCM, hypertrophic cardiomyopathy; CA, cardiac amyloidosis; AL-CA, AL cardiac amyloidosis; TTR-CA, transthyretin cardiac amyloidosis

### Hypertensive heart disease

Patients with a history of hypertension who met the diagnostic criteria for LVH on echocardiography (LV mass index [LVMI] >115 g/m^2^ for men, and >95 g/m^2^ for women), without other potential causes of LVH.(7)

### Hypertrophic cardiomyopathy

Patients who met the diagnostic criteria of HCM (LVWTmax ≥15 mm on echocardiography, in the absence of abnormal loading conditions that could sufficiently explain the LVH) were included.(8, 9) Definite evidence of HCM on CMR or identification of a typical gene mutation was required for a precise diagnosis.

### Cardiac amyloidosis

The types of CA were determined by evaluating serum and urine electrophoresis, serum free light chains, CMR imaging, and technetium-99m scintigraphy, and were confirmed based on the findings of myocardial biopsies.(5) AL-CA was defined as biopsy-proven AL amyloidosis and/or monoclonal gammopathy, accompanied by the following findings: (i) increased LV wall thickness without dilated LV (average LV wall thickness ≥12 mm) in the presence of low voltage QRS (amplitude <0.5 mV in the limb leads or <1.0 mV in the anterior leads); or (ii) typical findings on CMR (patchy, subendocardial circumferential, or diffuse fuzzy LGE of the LV). Patients who showed positive or equivocal TTR immunohistochemical staining on myocardial biopsy underwent direct screening analysis for detection of *TTR* mutations.(10)

### Fabry disease

FD was diagnosed based on the assessment of α-galactosidase A activity in the serum (for male patients) and the detection of *GLA* mutation (for both male and female patients).(11)

### Exclusion criteria

Patients with unknown etiology of LVH even after performing all available diagnostic tests were excluded. In addition, patients were excluded if they presented with any of the following: (1) significant LV dysfunction (LV ejection fraction <40%), (2) active malignancy (other than AL-CA) or undergoing chemotherapy, (3) end-stage renal disease, (4) prior coronary revascularization, (5) evidence of combined ischemic cardiomyopathy, (6) significant valve disease, (7) AA amyloidosis, or (8) other metabolic or infiltrative cardiomyopathies.

### Clinical data acquisition

Clinical and anthropometric, laboratory, and echocardiographic measurements were obtained at the time of CMR imaging. All echocardiographic images were obtained using a standard ultrasound device with a 2.5-MHz probe, in accordance with the guidelines of the European Association of Cardiovascular Imaging.(7)

### CMR imaging acquisition and assessment of myocardial fibrosis

Detailed procedure used for CMR imaging is provided in the **Supplementary Material**. CMR imaging was performed according to standard protocols using a 3.0-T imager (Ingenia CX, Philips Healthcare, Best, Netherlands) with a 16-channel phased array coil. Cine imaging was obtained using a segmented steady-state free-precession sequence. LGE images were obtained using a phase-sensitive inversion recovery sequence 10 min after the injection of 0.2 mmol/kg of gadobutrol (Gadovist, Bayer Schering Pharma, Berlin, Germany). Pre- and post-contrast (15 minutes) T1 mapping was performed using a mid-ventricular short-axis section at the level of papillary muscles, and the images were acquired using the modified Look-Locker inversion-recovery (MOLLI) sequence (the “3-3-5” standard protocol).(6) The region of interest was delineated on the entire LV myocardium at the level of papillary muscles.(12, 13) T1 maps were generated using MR workstation with in-line motion correction following image acquisition. The recovery rate of T1 relaxation was measured in a mid-ventricular short-axis slice.(13, 14) ECV was calculated as ECV = (1–hematocrit) × [△R1_myocardium_]/[△R1_blood_], with hematocrit measured at the time of CMR imaging.(6) All measurements were performed by a single radiologist (E-J.C.).

### AI-based assessment of native T1 and ECV

Our deep learning (DL) algorithm was used for automated analysis of T1 maps in accordance with the method described in the previous study,^19^ in which 2D U-Net for myocardial segmentation incorporated into Myomics-T1 software ver. 1.0.0 (Phantomics Inc.). Details of the architecture of the DL model are presented in the **Supplementary Materials** and **Figure S1**. Additional relevant information can be found in previous publications.(13, 14) Representative figures of the AI-derived native T1 and ECV for various etiologies of LVH are shown in **Figure 2**. The correlation and agreement analyses conducted using 30 randomly selected patients are summarized in **Figure S2**.

**Figure 2.**
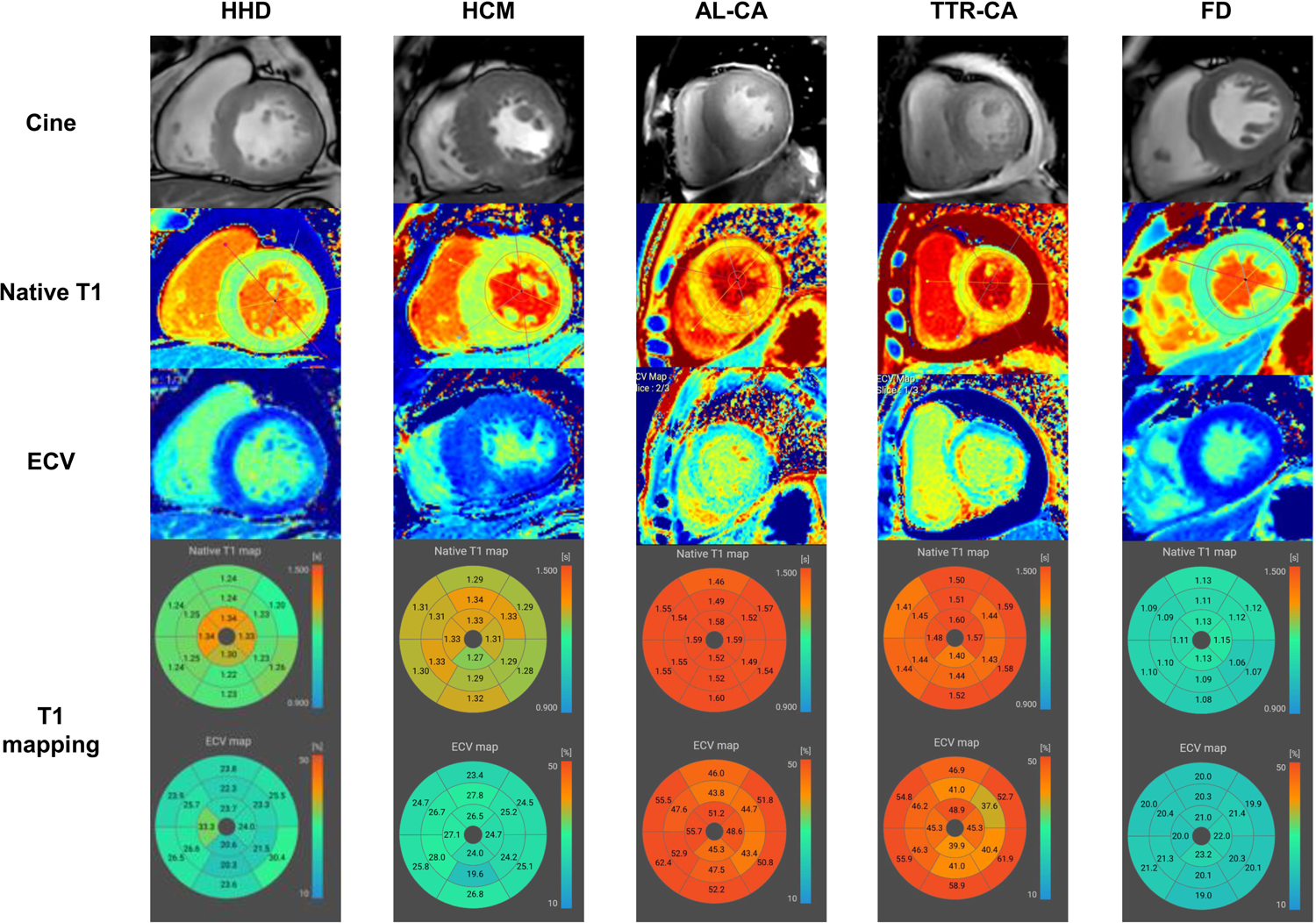
Representative figures of AI-derived native T1 and ECV. Typical native T1 and ECV measurements obtained through an AI-assisted semi-automated algorithm for various etiologies of LVH: (A) HHD, (B) HCM, (C) AL-CA, (D) TTR-CA, and (E) FD. Abbreviations: FD, Fabry disease; others as in Figure 1.

### Study outcomes

The diagnostic outcome was the area under the receiver operating characteristic curve (AUC) for differentiating CA from other etiologies of LVH. The clinical outcome for prognosis of AL-CA was a composite of cardiovascular death and hospitalization for heart failure, assessed through regular outpatient visits, telephone interviews and chart reviews.

### Statistical analysis

Categorical variables are presented as frequencies and percentages, and continuous variables as means ± standard deviations. Group comparisons were performed with Mann-Whitney U test and Kruskal-Wallis test for continuous variables, and the χ2 test or Fisher’s exact test for categorical variables. The AUC was used to measure the performance of the DL algorithm in the diagnosis of CA. Kaplan–Meier method and Cox proportional hazard regression model were used for survival analysis. Multivariate Cox proportional hazards regression with backward selection method was performed using univariable markers with p-values <0.100. All statistical analyses were performed using R statistical software version 3.6.3 (The R Foundation, Vienna, Austria), and p-value <0.05 was considered statistically significant.

## Results

### Baseline characteristics

Baseline characteristics of the study population are summarized in **Table 1**. The mean LVMI was 118.7±33.7 g/m^2^ in patients with HHD, 134.9±32.7 g/m^2^ in those with HCM, 134.0±36.1 g/m^2^ in those with AL-CA, 143.0±54.0 g/m^2^ in those with TTR-CA, and 162.2±80.2 g/m^2^ in those with FD. Patients with CA had the highest native T1 values (1444.6±83.4 msec in AL-CA, and 1418.3±72.8 msec in TTR-CA), followed by patients with HCM (1319.4±56.9 msec), and those with HHD (1291.4±38.9 msec), and patients with FD (1143.9±80.3 msec) (**Figure 3**). The ECV values showed similar trends: the highest in patients with AL-CA (43.6±8.1%), followed by those with TTR-CA (40.0±4.3%), HCM (29.5±5.9%), and HHD (26.1±3.3%), and FD (23.2±3.0%) (**Figure 3**).

**Figure 3.**
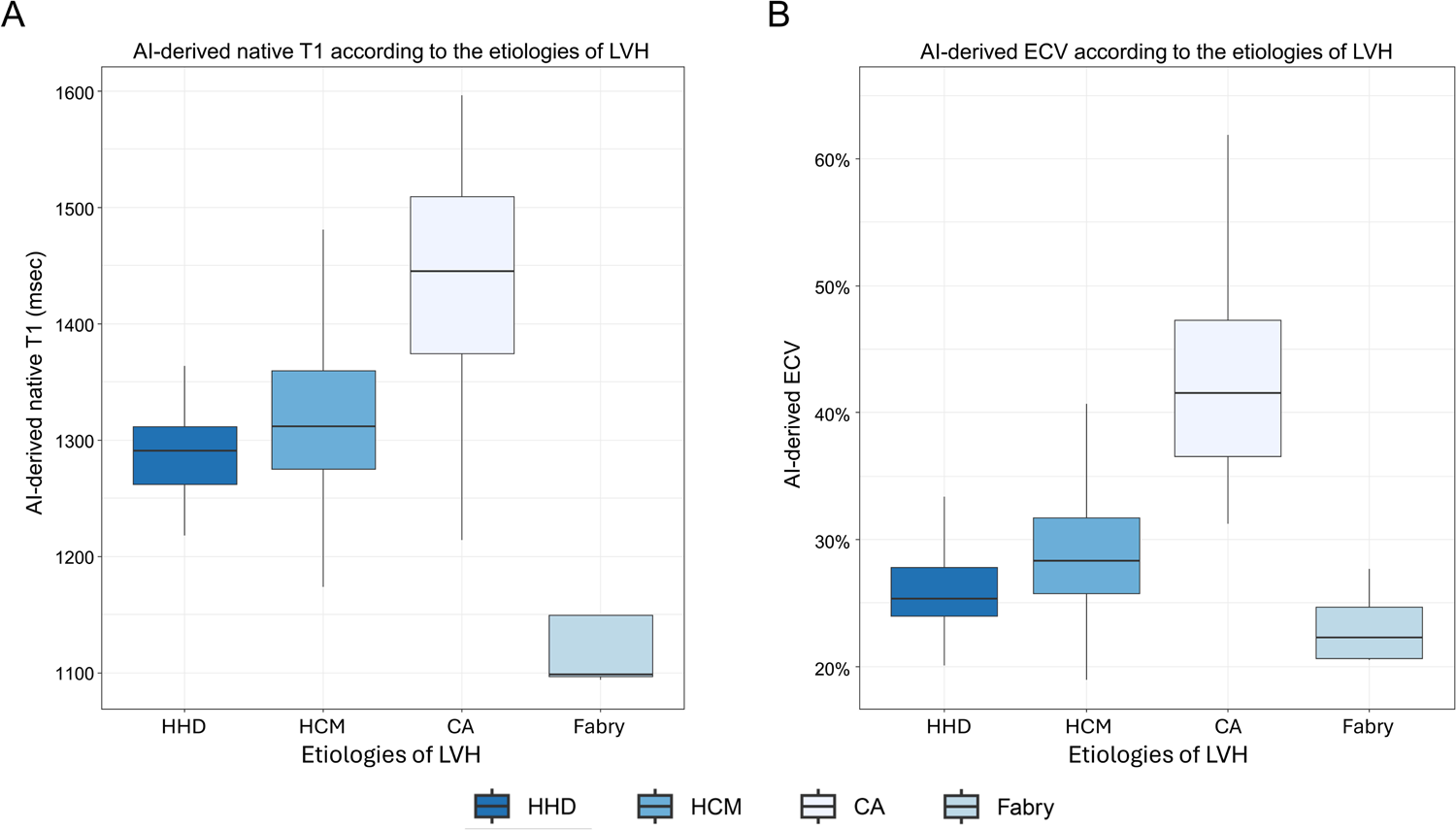
Native T1 and ECV measurements according to the LVH etiologies. AI-derived native T1 (A) and ECV (B) measurements are shown across various etiologies of LVH. Abbreviations as in Figure 1.

**Table 1.** Baseline characteristics.

|  | **HHD** | **HCM** | **CA (n=50)** |  | **Fabry disease** | **p-value** |
| --- | --- | --- | --- | --- | --- | --- |
|  | **(n=47)** | **(n=198)** | **AL-CA (n=39)** | **TTR-CA (n=11)** | **(n=5)** |  |
| **Age, years** | 56.4±16.3 | 57.6±13.8 | 69.8±10.4 | 78.7±5.3 | 45.9±3.3 | <0.001 |
| **Male** | 30 (63.8%) | 143 (72.2%) | 24 (61.5%) | 3 (27.3%) | 4 (80.0%) | 0.022 |
| **Body-mass index, kg/m<sup>2</sup>** | 24.8±3.6 | 25.7±3.9 | 24.2±2.7 | 21.9±9.2 | 22.5±2.2 | 0.028 |
| **Systolic blood pressure, mmHg** | 136.8±22.8 | 130.3±15.9 | 115.4±20.4 | 128.5±23.3 | 140.8±12.5 | <0.001 |
| **Diastolic blood pressure, mmHg** | 81.9±15.4 | 76.2±12.2 | 70.4±12.7 | 68.5±4.9 | 79.8±7.5 | 0.001 |
| **Hypertension** | 47 (100.0%) | 66 (33.3%) | 13 (33.3%) | 1 (9.1%) | 0 (0.0%) | 0.129 |
| **Diabetes mellitus** | 13 (27.7%) | 49 (24.7%) | 10 (25.6%) | 1 (9.1%) | 0 (0.0%) | 0.641 |
| **Chronic kidney disease** | 5 (10.6%) | 6 (3.0%) | 13 (33.3%) | 0 (0.0%) | 2 (40.0%) | <0.001 |
| **Hemoglobin, g/dL** | 13.7±2.3 | 14.4±1.8 | 11.8±1.6 | 10.7 ± 0.6 | 13.2 ± 1.0 | <0.001 |
| **Hematocrit, %** | 40.8±6.3 | 42.9±5.1 | 35.7±4.6 | 34.0±0.2 | 40.2±3.1 | <0.001 |
| **Serum creatinine, mg/dL** | 1.31±1.81 | 0.87±0.22 | 1.02±0.43 | 0.99±0.52 | 0.97±0.19 | 0.015 |
| **GFR, mL/min/1.73m<sup>2</sup>** | 80.9±27.5 | 90.1±16.7 | 73.3±25.6 | 63.0±39.6 | 89.0±12.6 | <0.001 |
| **CKMB, ng/mL** | 6.2±10.5 | 11.7±49.1 | 4.6±3.2 | 2.9±2.8 | 5.7±1.8 | 0.901 |
| **Troponin I, ng/mL** | 1.05±3.22 | 1.65±5.40 | 0.48±0.73 | 0.04±0.00 | 0.34±0.59 | 0.664 |
| **NT-proBNP, pg/mL** | 1391.0±3055.5 | 1832.2±2383.8 | 8023.6±6639.1 | 1327.9±1119.5 | 1670.2±2778.8 | <0.001 |
| **Echocardiographic parameters** |  |  |  |  |  |  |
| **LV-EDV, mL** | 83.6±27.9 | 77.4±25.9 | 67.6±19.1 | 64.5±12.0 | 95.8±19.5 | 0.021 |
| **LV-ESV, mL** | 34.3±18.3 | 28.2±13.1 | 30.3±11.8 | 31.0±22.6 | 29.6±6.7 | 0.125 |
| **LV-EF, %** | 60.3±9.5 | 63.7±7.2 | 55.6±8.8 | 54.4±26.6 | 69.2±1.9 | <0.001 |
| **LVMI, g/m<sup>2</sup>** | 118.7±33.7 | 134.9±32.7 | 134.0±36.1 | 143.0±54.0 | 162.2±80.2 | 0.017 |
| **LAVI, mL/m<sup>2</sup>** | 42.6±16.4 | 50.7±19.5 | 54.8±14.6 | 63.0±22.5 | 44.0±11.1 | 0.020 |
| **E/e' ratio** | 13.5±6.8 | 14.6±6.8 | 26.0±8.9 | 28.5±8.9 | 17.6±2.7 | <0.001 |
| **TR Vmax, m/sec** | 2.5±0.4 | 2.4±0.3 | 4.2±8.1 | 3.3±0.1 | 2.5±0.5 | 0.034 |
| **LV-GLS, %** | 13.9±2.9 | 11.9±3.8 | 9.5±3.4 | 14.8±4.0 | 11.5±5.0 | <0.001 |
| **CMR measurements** |  |  |  |  |  |  |
| **Native T1, msec** | 1291.4±38.9 | 1319.4±56.9 | 1444.6±83.4 | 1418.3±72.8 | 1143.9±80.3 | <0.001 |
| **Post T1, msec** | 486.1±47.2 | 451.2±71.9 | 397.3±79.9 | 365.3±73.9 | 496.9±57.1 | <0.001 |
| **Post-contrast T2, msec** | 49.6±4.0 | 49.5±3.1 | 61.6±11.6 | 52.3±2.8 | 46.8±4.8 | <0.001 |
| **ECV, %** | 26.1±3.3 | 29.5±5.9 | 43.6±8.1 | 40.0±4.3 | 23.2±3.0 | <0.001 |

Values are presented as the mean±standard deviation or as a number (percentage).
Abbreviations: BMI, body-mass index; GFR, glomerular filtration rate; CKMB, creatine kinase-myocardial band; NT-proBNP, N-terminal pro-B natriuretic peptide; LV, left ventricular; EDV, end-diastolic volume; ESV, end-systolic volume; EF, ejection fraction; LAVI, left atrial volume index; LVMI, left ventricular mass index; TR, tricuspid regurgitation; LV-GLS, left ventricular global longitudinal strain; ECV, extracellular volume fraction; CMR, cardiac magnetic resonance.

### Accuracy of AI-derived native T1 and ECV measurements

The AI-derived and manual native T1 and ECV measurements were compared on a per-patient basis (**Figure S2**). AI-derived native T1 showed a strong correlation and good agreement with the reference native T1 (r=0.995, p<0.001; bias 2.3 msec, 95% limits of agreement [LoA] −15.0 to 19.6 msec). In addition, AI-derived ECV was strongly correlated with the reference ECV (r=0.993, p<0.001) with a good agreement (bias 0.2%, 95% LoA −2.2% to 2.6%).

### Differentiation of CA from other etiologies of LVH

The diagnostic performance of the AI-derived native T1 and ECV measurements was assessed in terms of differentiating CA from other etiologies of LVH. The AUC values for the diagnosis of CA were 0.899 (95% CI 0.847–0.952; p<0.001) for native T1 and 0.946 (95% CI 0.922–0.971; p<0.001) for ECV (**Figure S3**). The optimal native T1 and ECV cutoff values were 1364 msec and 33.6%, respectively (accuracy, 84.0% and 85.6%; sensitivity, 84.5% and 87.5%; and specificity, 86.8% and 96.2%, respectively).

### Prognostication with native T1 and ECV for AL-CA

Thirty-nine patients in the study population were diagnosed with AL-CA: 14 (35.9%) were at revised Mayo stage III and 25 (64.1%) were at stage IV. During a mean 22 months of follow-up, 17 patients died of cardiovascular causes and 20 patients were hospitalized for heart failure. The AUC values for the prediction of study outcome using native T1 and ECV were 0.658 (95% CI 0.462–0.855; p=0.139) and 0.888 (95% CI 0.764–1.000; p<0.001), respectively (p-for-comparison=0.05; **Figure S4**). The optimal cutoff value of ECV for study outcome was 40.0% (accuracy, 92.9%; sensitivity, 79.0%; and specificity, 70.8%).

Survival curves of patients with AL-CA are depicted in **Figure 4**, according to the difference in the serum free light chain (dFLC) (**Figure 4A**), NT-proBNP (**Figure 4B**), revised Mayo stage (**Figure 4C**), and AI-derived ECV (**Figure 4D**). The AI-derived ECV showed a significant discrimination of prognosis (adjusted HR 4.247 for ECV ≥40%, 95% CI 1.215–14.851, p=0.024), whereas other conventional risk parameters, such as the dFLC, NT-proBNP, cardiac troponin, and LV global longitudinal strain, did not show significant prognostic value (**Table 2** and **Table S1**). The prognostic value of the AI-derived ECV remained significant even after adjusting for the revised Mayo stage (adjusted HR 6.324 for ECV ≥40%, 95% CI 1.794–22.297, p=0.004).

**Figure 4.**
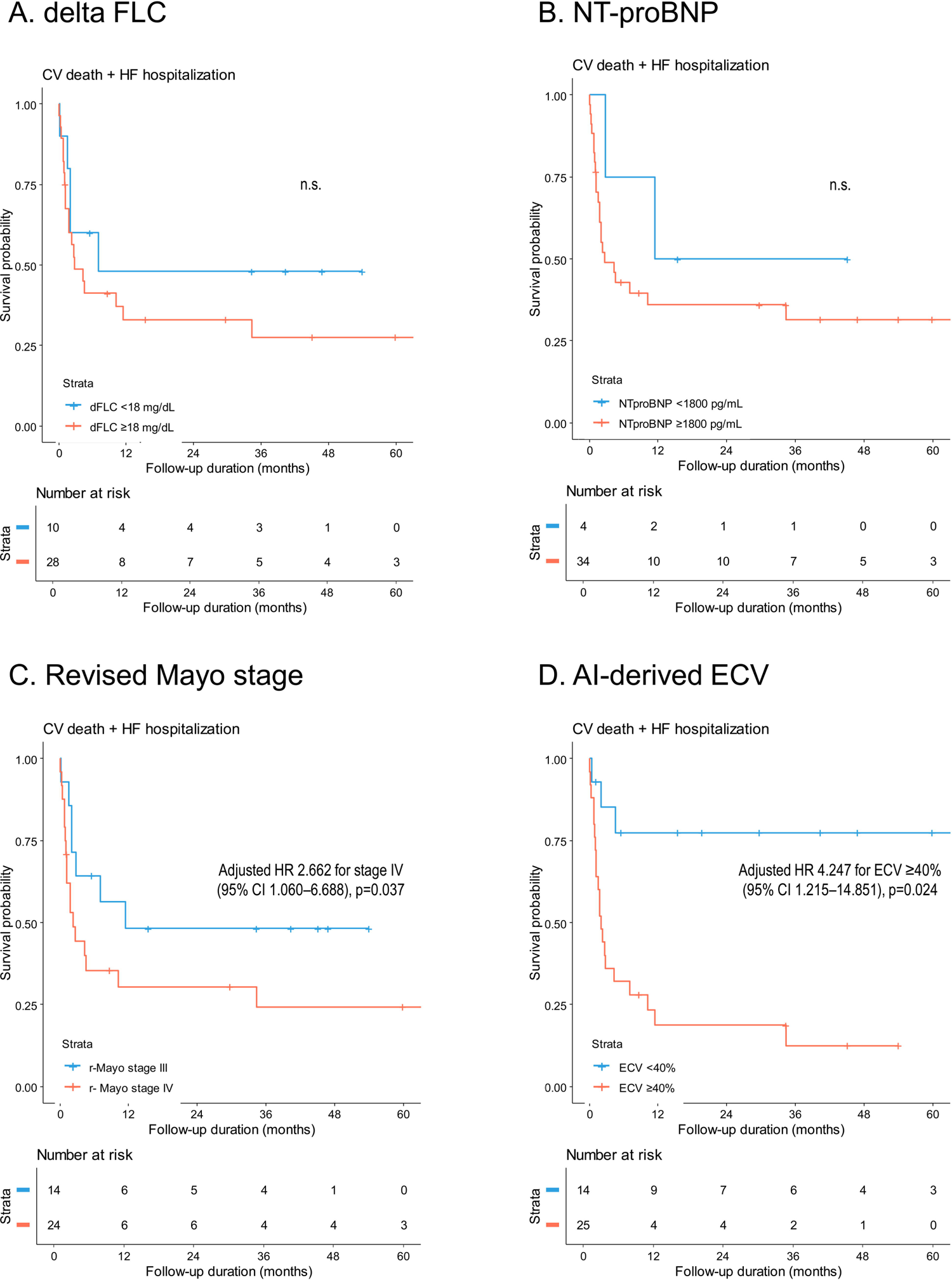
Event-free survival of patients with AL-CA. Clinical outcomes were compared between the subgroups divided by (A) dFLC, (B) NT-proBNP, (C) revised Mayo stage, and (D) AI-derived ECV. Abbreviations: dFLC, difference in the serum levels of free light chain; NT-proBNP, N-terminal pro-B natriuretic peptide; n.s., not significant; others as in Figure 1.

**Table 2.** Multivariable predictors of composite outcome among patients with AL cardiac amyloidosis.

|  | Multivariable analysis<br>using continuous variables |  | Multivariable analysis<br>using categorical variables |  | Multivariable analysis<br>using revised Mayo stages<br>combined with ECV |  |
| --- | --- | --- | --- | --- | --- | --- |
|  | Adjusted HR (95% CI) | P-value | Adjusted HR (95% CI) | P-value | Adjusted HR (95% CI) | P-value |
| **SBP (per +1 mmHg)** | 0.963 (0.935–0.992) | 0.014 |  |  |  |  |
| **SBP <110 mmHg** |  |  | 2.575 (1.074–6.173) | 0.034 | 1.478 (0.493–4.435) | 0.485 |
| **NT-proBNP (per +1 pg/mL)** | 1.000 (1.000–1.000) | 0.847 |  |  |  |  |
| **Troponin T ≥0.025 ng/mL** |  |  | 1.039 (0.372–2.904) | 0.942 |  |  |
| **LVGLS (per +1%)** | 0.850 (0.703–1.027) | 0.092 |  |  |  |  |
| **LVGLS <8%** |  |  | 1.888 (0.787–4.525) | 0.154 |  |  |
| **LVGLS <10%** |  |  |  |  | 2.116 (0.806–5.559) | 0.128 |
| **Revised Mayo stage** |  |  |  |  | 2.662 (1.060–6.688) | 0.037 |
| **ECV (per +1%)** | 1.080 (1.011–1.153) | 0.023 |  |  |  |  |
| **ECV ≥40%** |  |  | 4.247 (1.215–14.851) | 0.024 | 6.324 (1.794–22.297) | 0.004 |

Abbreviations: HR, hazard ratio; CI, confidence interval; others as in Table 1.

The prognostic value of the AI-derived ECV in patients with AL-CA was further assessed using the following subgroups: group 1 (revised Mayo stage III and ECV <40%), group 2 (revised Mayo stage IV and ECV <40%), group 3 (revised Mayo stage III and ECV ≥40%), and group 4 (revised Mayo stage IV and ECV ≥40%). The combination of revised Mayo stage and AI-derived ECV showed excellent risk stratification, with the worse prognosis in group 4 and the best in group 1 (**Figure 5**). The AUC of the combination of revised Mayo stage and AI-derived ECV was significantly higher than that of the Mayo staging system alone (AUC 0.860 vs. 0.588; p=0.006). In addition, net reclassification improvement (NRI) and integrated discrimination improvement (IDI) analyses showed that the combination of ECV and the revised Mayo staging system could provide significantly better risk stratification than the revised Mayo staging system alone (IDI 0.279, 95% CI 0.025–0.499, p=0.013; NRI 0.138, 95% CI 0.000–0.597, p=0.007; **Table 3**).

**Figure 5.**
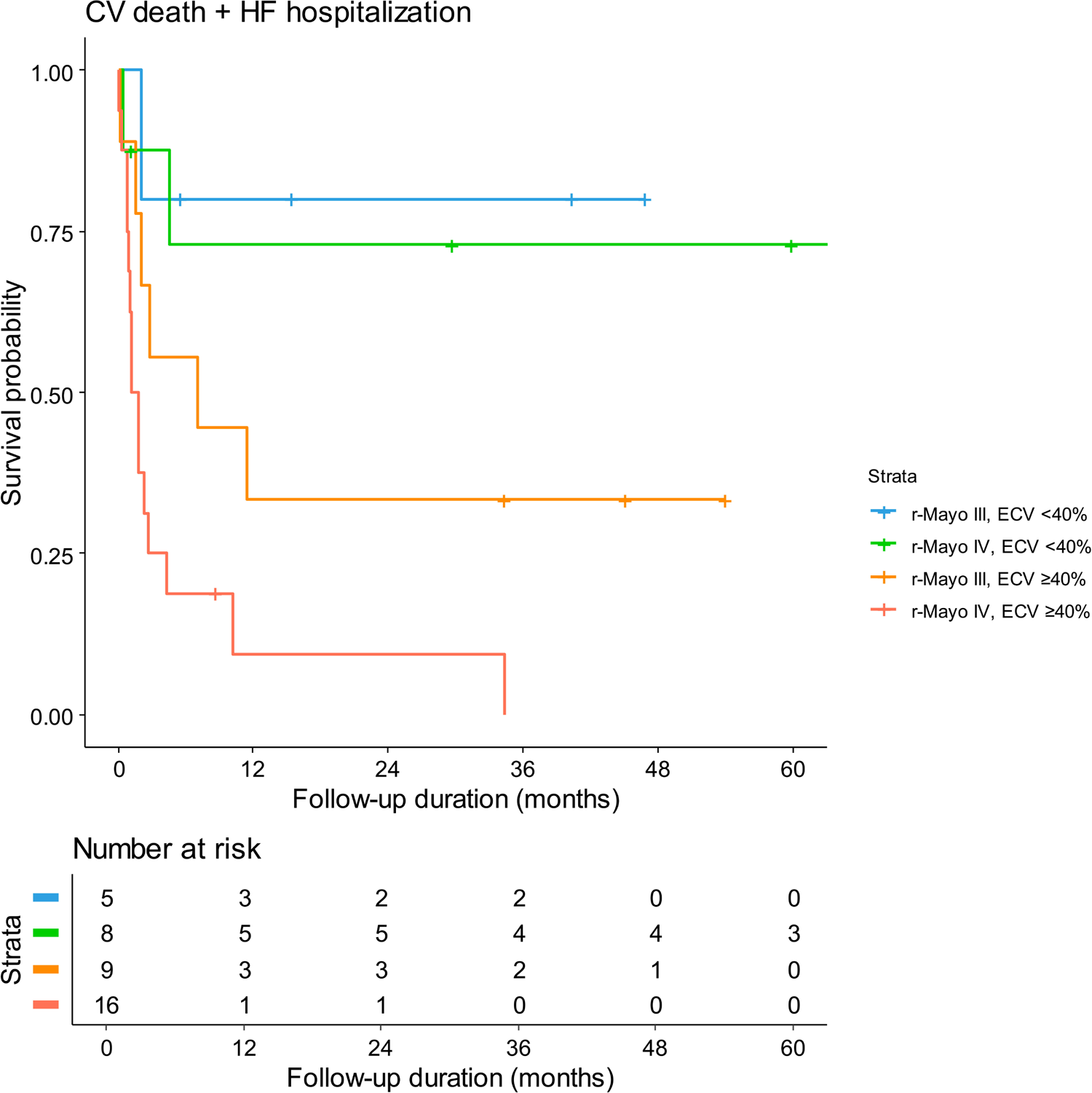
Event-free survival of patients with AL-CA according to the combination of revised Mayo stage and AI-derived ECV measurement. Clinical outcomes were compared between the subgroups divided by a combination of revised Mayo staging and AI-derived ECV. Abbreviations as in Figure 1.

**Table 3.** Incremental prognostic value of AI-derived ECV over revised Mayo staging.

|  | Value | 95% CI | p-value |
| --- | --- | --- | --- |
| IDI (combination of revised Mayo stage and ECV vs. revised Mayo stage only) | 0.279 | 0.025–0.499 | 0.013 |
| NRI (combination of revised Mayo stage and ECV vs. revised Mayo stage only) | 0.138 | 0.000–0.597 | 0.007 |
| AUC of the combination of revised Mayo stage and ECV | 0.860 | 0.740–0.979 | 0.006 |
| AUC of revised Mayo stage | 0.588 | 0.427–0.748 |  |

Abbreviations: AUC, area under the curve; IDI, integrated discrimination improvement; NRI, net reclassification improvement.

## Discussion

In this study, we analyzed the performance of AI-derived native T1 and ECV in the diagnosis and prognostication of CA. The results demonstrated that the AI-derived native T1 and ECV were effective in differentiating CA from other etiologies of LVH. Furthermore, in patients with AL-CA, the AI-derived ECV showed independent prognostic value beyond the revised Mayo stage and could provide better risk stratification when incorporated into current staging system. Our findings suggest the usefulness of AI-derived T1 mapping for differential diagnosis of LVH, as well as for prognostication in AL-CA.

### Etiologies of LVH and the importance of differential diagnosis

LVH is a common cardiac condition, but various etiologies should be considered because of the considerable differences in the pathophysiology, treatment, and prognosis across the possible etiologies of LVH.(1, 3) If LVH is a result from increased afterload, i.e., HHD, management is mainly focused on the alleviation of blood pressure, and the prognosis is benign. However, in patients with HCM, the assessment of sudden cardiac death risk and its prevention is crucial in the treatment strategy, with special considerations required for its complications, such as arrhythmia, heart failure, or LV outflow tract obstruction.(8, 9) AL-CA requires a complex approach, given that AL-CA is a hematologic malignancy with excessive production of light-chain immunoglobulins.(2) Its management includes cytotoxic chemotherapy and stem cell transplantation, along with the management of cardiovascular complications, but the prognosis is poor: patients with AL-CA have a median survival of 24 months from the initial diagnosis.(5, 15) As a subtype of CA, TTR-CA shares common morphological features with AL-CA; however, its pathophysiology is unique. The accumulation of TTR, due to genetic or unknown causes, is the main target for specialized treatment (i.e., TTR tetramer stabilizers).(10) Another unique etiology of diffuse LVH should be noted: FD is a rare X-linked lysosomal storage disorder, caused by mutations in *GLA*, responsible for coding the lysosomal enzyme α-galactosidase A.(11) The absent or insufficient α-galactosidase A activity leads to the accumulation of globotriaosylceramide and related globotriaosylsphingosine in affected tissues. This accumulation in myocardium results in myocardial fibrosis, requiring specific therapies, primarily enzyme replacement therapy.

### Differential diagnosis of LVH etiologies using CMR

The morphological similarities of different etiologies of LVH and the lack of pathognomonic/specific findings makes its differential diagnosis using echocardiography challenging.(3) Therefore, an arbitrary diagnosis based on echocardiography needs to be investigated further using non-invasive imaging modalities, such as CMR. In particular, HCM typically shows multifocal discrete LGE, CA shows diffuse LGE typically at subendocardial layer, FD shows diffuse LGE across entire myocardium, and HHD shows focal fuzzy LGE at the right ventricular insertion points of LV myocardium.(12) However, these patterns can overlap between etiologies, and may not even be apparent in the early stages of the disease. Given these limitations and ambiguity in the assessment of LGE patterns for the differential diagnosis of LVH etiologies, several studies have been conducted to analyze the efficacy of quantitative assessment using native T1 and ECV.(6, 16) Native T1 and ECV typically reflect the degree and amount of myocardial fibrosis, not only in the advanced stage (so called “replacement fibrosis”) but also in the early stage of diffuse myocardial fibrosis.(17) The accumulation of robust evidence has led to steady evolution of the use of native T1 and ECV in myocardial disease, showing different patterns of native T1 and ECV between the etiologies of LVH. According to a study by Baggiano et al., native T1 enabled diagnosis of CA, with a cutoff native T1 of <1036 msec resulting in a 98% negative predictive value and native T1 value of >1164 msec resulting in a 98% positive predictive value.(18) Hinojar et al reported that patients with HCM show significantly higher native T1 and ECV than those with HHD.(19) FD is also known to have significantly lower native T1 values than HHD and HCM.(20, 21) Our findings are consistent with previous studies, showing that CA results in the highest native T1 and ECV, followed by HCM and HHD, and with FD showing the lowest values.

Although both native T1 and ECV can be useful in differential diagnosis of LVH etiologies, it should be noted that native T1 can be affected by MR vendors, the intensity of magnetic field, and the amount and concentration of contrast agent. On the other hand, ECV represents a physiological parameter and is derived from the ratio of T1 signal values, therefore could be more reproducible between different field strengths, vendors, and acquisition techniques than both native and post-contrast T1.(16) ECV measures also exhibit better agreement with histological measures of the collagen volume fraction than isolated post-contrast T1. Given the better performance of ECV (AUC 0.946) in differentiating CA from other etiologies of LVH than that of native T1 (AUC 0.899), our findings support that ECV measurements could be more relevant than native T1 values for diagnosis of CA.

### Deep learning algorithm for the measurement of native T1 and ECV

A previous study of 95 participants, including 12 patients with HCM, 12 with CA, and 12 with FD, conducted using our DL model demonstrated excellent correlation and agreement between the AI-derived native T1 and ECV and the reference values on a per-patient basis (native T1: r=0.967, bias 9.5 msec; ECV: r=0.987, bias 0.7%).(13) The excellent accuracy of our DL algorithm, which was further confirmed in the present study, indicates that the use of DL algorithm on the T1 mapping images can considerably reduce the time required for image interpretation.(13) In addition, considering that the differential diagnosis of LVH requires a series of non-invasive and invasive tests, the accuracy of T1 mapping parameters in the detection of CA can improve the diagnostic process in clinical practice. In particular, the differential diagnosis process includes myocardial biopsy for confirming amyloid infiltration in myocardium with specific immunohistochemistry staining for determination of CA subtypes. Genetic tests are required for the confirmation of HCM and TTR-CA, a process that leads to some delays until the final diagnosis. Thus, accurate interpretation of CMR images using T1 mapping can focus down to the possible diagnosis and facilitate the selection of appropriate diagnostic tests.

### Prediction of prognosis in AL-CA using T1 mapping parameters

In the present study, we assessed the prognostic value of ECV, because in addition to ECV having theoretical advantages over native T1, the AUC of ECV in the present study was higher than that of native T1, not only for differentiating CA from other LVH etiologies but also for predicting its prognosis. Additionally, because of the small number of events in patients with HHD and HCM, and the small number of patients with TTR-CA and FD, we focused our analysis on patients with AL-CA.

Several clinical tools have been used to assess the prognosis of patients with AL-CA. The revised Mayo staging system suggested in 2012, which is the most widely used clinical staging system for AL-CA, incorporates multiparametric biomarkers, such as the levels of NT-proBNP, cardiac troponin, and the dFLC.(22) However, it could be argued that the incorporation of myocardial status into the system would be more representative of the disease stage, because the most common cause of death in patients with AL-CA is cardiovascular death. Indeed, the incorporation of LGE on CMR image into the staging system provided promising results.(23) However, it should be noted that the determination of LGE in AL-CA is often obscure, due to the fuzzy involvement of amyloid. Instead, the T1 mapping could be promising, given its more feasible quantitation and direct reflection of the burden of amyloid infiltration in myocardium.(24) Moreover, patients with revised Mayo stage III and IV in the present study who had elevated ECV (indicative of advanced myocardial infiltration) demonstrated higher risks of mortality than those with lower ECV. Furthermore, ECV had a more significant predictive value than the components of the revised Mayo staging system, such as the levels of NT-proBNP, cardiac troponin, and dFLC. These findings suggest that direct assessment of myocardial fibrosis using T1 mapping techniques could be more suitable for the prediction of mortality risk than indirect measurement of amyloid infiltration using serum biomarkers. Additionally, given that this is the first study to demonstrate the relevance of DL model not only for the diagnosis of CA, but also for prognostication in patients with AL-CA, our findings will facilitate the clinical application of DL models in CMR imaging.

### Limitations

The present study has certain limitations. First, this was a single-center retrospective study with a limited sample size. Second, the prognostic value of T1 mapping parameters was assessed only among the patients with AL-CA, but not for those with other etiologies. Patients with HCM and HHD did not experience sufficient number of events, and the number of patients with FD was too small for further analysis. Third, data on the quantitative measurement of LGE was not available for our study population. Instead, we focused on the clinical usefulness of AI-derived T1 mapping parameters. Nonetheless, we acknowledge that future studies on the radiomics features of LGE in the AI-derived assessment of CMR images are warranted. Fourth, our study focused on the clinical utilization of AI-derived measurements of T1 mapping parameters, rather than the automatization of the entire clinical process. Finally, the findings of the present study should be validated further in terms of improving the diagnostic process (total duration and costs for confirmative diagnosis) and prognosis.

## Conclusion

This study demonstrated that AI-derived native T1 and ECV on CMR images could differentiate CA from other etiologies of LVH. Furthermore, this study showed that AI-derived ECV has significant prognostic value in patients with AL-CA, suggesting its usefulness in clinical practice.

## Data Availability

Data used in this study cannot be made publicly available because of the strict ethical restrictions set by the IRB of Seoul National University Bundang Hospital (https://e-irb.snubh.org). Please contact the corresponding authors or the ethics board at SNUBH for further inquiries regarding data availability within the scope permitted by the IRB.

## Acknowledgements

None.

