## Supplementary materials for "Artificial Intelligence-Derived Extracellular Volume Fraction for Diagnosis and Prognostication in Patients with Light-Chain Cardiac Amyloidosis"

**Short title:** AI-derived T1 mapping for cardiac amyloidosis

**I. Supplementary Methods	p.2**

**II. Supplementary Figures	p.7**

**III. Supplemental Table	p.11**

**IV. References	p.13**

**I. Supplementary Methods**

***1. CMR imaging***

Cardiac magnetic resonance (CMR) imaging was conducted in accordance with standard protocols using a 3.0-T imager (Ingenia CX, Philips Healthcare, Best, Netherlands) equipped with 16-channel phased-array coils.(1, 2) Balanced steady-state free-precession cine images, including short-axis, four-chamber, and two-chamber images, were obtained during breath holding. Left ventricular (LV) short-axis images were acquired at 10-mm intervals (6-mm thickness with 4-mm intersection gap), encompassing the entire LV volume from the base to apex, using retrospective electrocardiographic gating with the following parameters: repetition time (TR)/echo time (TE), 2.8–3.2 msec/1.4–1.6 msec; flip angle, 50°; 20 phases per cardiac cycle and nine k-space lines/segment; field of view, 250×250 mm; and matrix, 124×121. The acceleration factor was 1.5 or 3.6 depending on the number of slices acquired per breath hold.

LV diastolic and systolic volumes, as well as LV mass, were calculated from the cine images and indexed for body-surface area. LV mass was measured by multiplying the sum of total LV myocardial volumes by the specific gravity of myocardium. Following the acquisition of the cine images, a midventricular short-axis section was obtained at the papillary muscle level using the modified Look-Locker inversion-recovery (MOLLI) sequence applied for both pre- and post-contrast T1 mapping, and a single-short bSSFP sequence with the following parameters: TR/TE, 2.2/1.01 msec, flip angle, 20°; field of view (FOV), 250×250 mm^2^; section thickness, 8 mm, and acceleration factor, 2. Subsequently, 11 images acquired during 17 heartbeats were obtained, and in-line motion correction and map generation were performed. Post-contrast T1 mapping images were produced at the same positions within 15 minutes after injecting the contrast agent.

Late gadolinium enhancement (LGE) images were obtained 10 minutes after intravenous administration of 0.2 mmol/kg of gadolinium (Gadovist, Bayer Schering Pharma; Berlin, Germany) using a phase-sensitive inversion-recovery (PSIR) turbo field echo sequence with the following parameters: slice thickness, 8 mm; interslice gap 2 mm; TR, 6.2 msec; TE, 3.0 msec; flip angle, 25 degrees; and FOV, 320×320 mm^2^. All LGE-CMR images were analyzed by an experienced radiologist (E-J.C.), who was blinded to the patient information. The region of myocardial fibrosis was delineated as the sum of pixels with a signal intensity exceeding 5 standard deviations (SD) of the normal remote myocardium at each short-axis slice, using an appropriate post-processing program (CVI42, Circle Cardiovascular Imaging Inc., Calgary, Canada).(3)

***2. Artificial intelligence-based assessment of native T1 and ECV***

We conducted an automated analysis of the T1 maps, using a deep learning (DL) algorithm based on 2D U-Net (Myomics-T1 software, version 1.0.0., Phantomics Inc.), according to the procedure described in previous studies.(4, 5) This algorithm provides automatic segmentation of the LV myocardium on the maps, as well as the native T1, post-T1, and ECV values in 16 segments according to the guidelines of the American Heart Association.(6) A 10% epicardial and endocardial offset was applied to include only myocardial tissue. The hematocrit level was entered directly by the user.

**Manual segmentation**

Manual annotation of the myocardium served as the ground truth for training the convolutional neural network (CNN) model and performing automatic segmentation. Manual annotation was conducted using commercial software (CVI42, Circle Cardiovascular Imaging Inc., Calgary, Canada). Specifically, two experienced researchers manually drew the endocardial and epicardial LV contours in the fixed cardiac phase, with the endocardial contour encompassing the papillary muscles and trabeculations. After the initial annotation, two cardiac radiologists meticulously reviewed all contours and rectified any inaccuracies.

After manual segmentation, a parsing procedure was employed to utilize the annotation as the ground truth for training and evaluation. The software outputted the indices of the contours in xml format. The x- and y-coordinates were converted into points in a 2D matrix. The exterior of the endocardial contour and the interior of the epicardial contour were then filled with 1. The final format of the annotation was a 2D mask image, in which the myocardium was labeled as 1.

***Pre-processing***

All datasets imported in DICOM format were subjected to a series of image-processing steps. To minimize variations in size, resolution, and signal intensity, the image processing steps included restoration of the resolution, cropping of the center region, and intensity normalization. If the images were interpolated to double their sizes in the reconstruction process, they were resized to the original matrix size, and a centered 128 × 128 region was cropped as the region of interest (ROI). After cropping the ROI, the signal intensity was normalized to the 0–1 range. The same pre-processing procedure was applied to the labeled images, with the myocardium assigned as 1 and the background as 0.

***Training the U-Net***

We used a well-established CNN network U-net architecture for myocardial segmentation on T1 maps. The detailed construction of the network for our study is described in **Figure 2**. Specifically, the 2D U-Net model was trained to predict the LV myocardium using respective data. The performance and generalization of the model were improved by applying data augmentation, including rotation, flipping, and shifting, to the training dataset. The models were constructed using the Keras deep learning library in TensorFlow (https://www.tensorflow.org/). The training and internal validation processes were implemented on an Ubuntu 18.04 system with Intel(R) Xeon Silver 4116 CPU @ 2.10 GHz and an Nvidia RTX 3090 GPU (24GB memory). The Adam optimizer with a learning rate = 1.0E–04, β1 = 0.9, β2 = 0.999, and batch size = 32 was used to compile the models. For the activation function, a rectified linear unit (ReLU) was used in the convolution layer, except for the last layer (softmax layer, simple binary thresholding). A Dice loss function was employed to train the models. The network was trained for 150 epochs, and it took 4–6 hours on a single GPU to train the network.

***Post-processing***

The trained model inferred the probability map of each pixel that estimated the location of the myocardium. The 2D predictions were rescaled to their original sizes and resolutions. If multiple disconnected components were present in the prediction images after the rescaling procedure, the largest component was considered to be the myocardial region and was stored in the images. The other isolated pixels were considered noise and were therefore removed.

***Evaluation of the model***

The performance of the trained models was evaluated by calculating the following Dice similarity coefficient (DSC):

$$DSC=\frac{2\left| X\cap Y \right|}{\left| X \right|+\left| Y \right|}$$

between the ground truth and the segmentation results. $X$ and $Y$ denote the given sets and $\left| X \right|$ denotes the number of elements in $X$. The results were generated using the internal validation dataset and the abovementioned model training and post-processing procedures. The DSC of each image was calculated using the results and ground truth. The mean DSC of the native T1 model was 0.87 and that of the post-contrast T1 was 0.78.

***Automatic analysis and reporting***

The measured myocardial T1 values were expressed as 16 American Heart Association segments in the form of a bull’s eye map. Three reference points (the center of mass of the LV endocardial contour and two right ventricular insertion points) were used to generate the bull’s eye map.

**II. Supplementary Figures**

**Figure S1. Schematic figure of the AI-derived native T1 and ECV measurements**

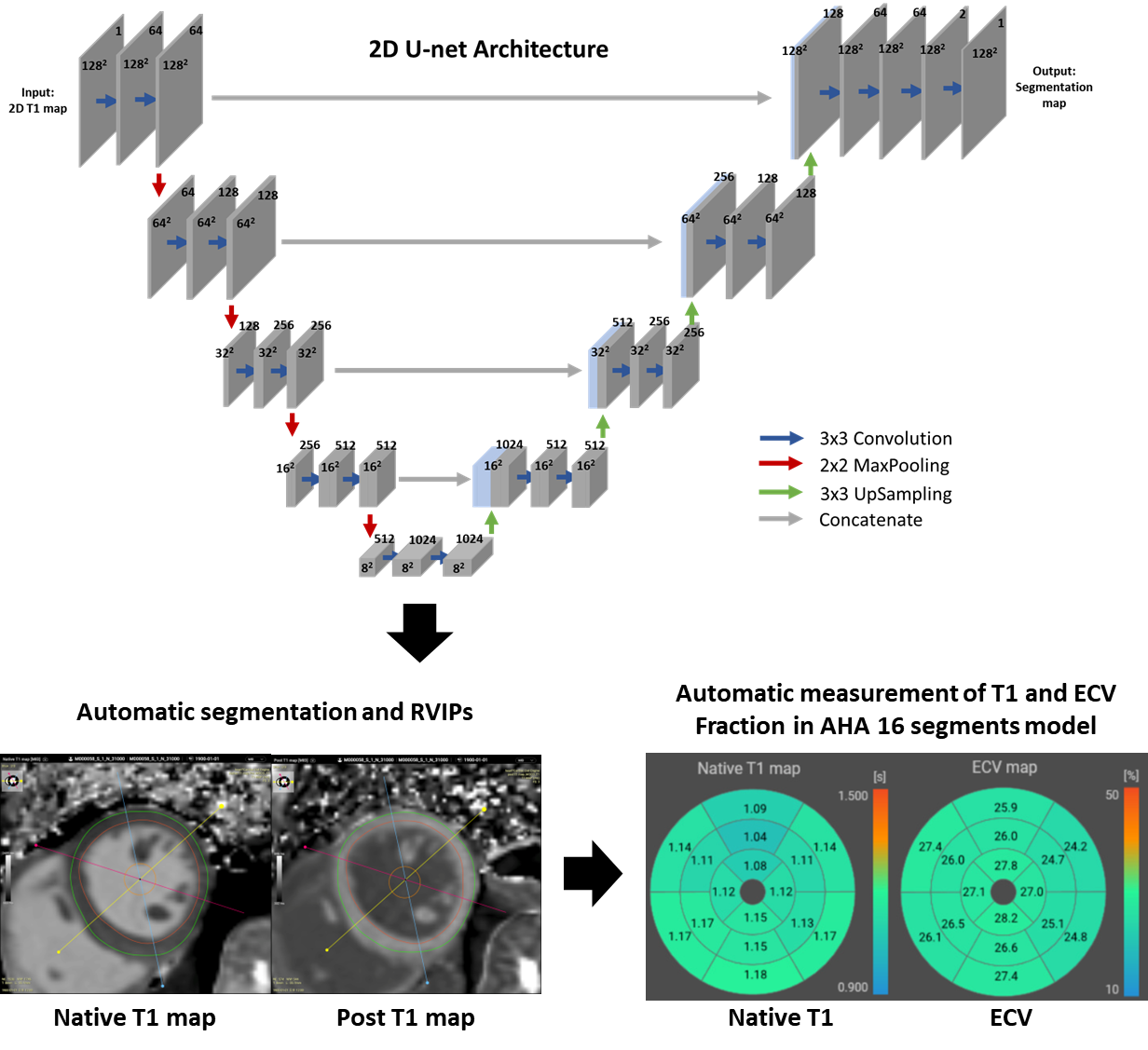

Abbreviations: RVIP, right ventricular insertion point; ECV, extracellular volume fraction; AHA, American Heart Association

**Figure S2. Accuracy of AI-derived native T1 and ECV measurements**

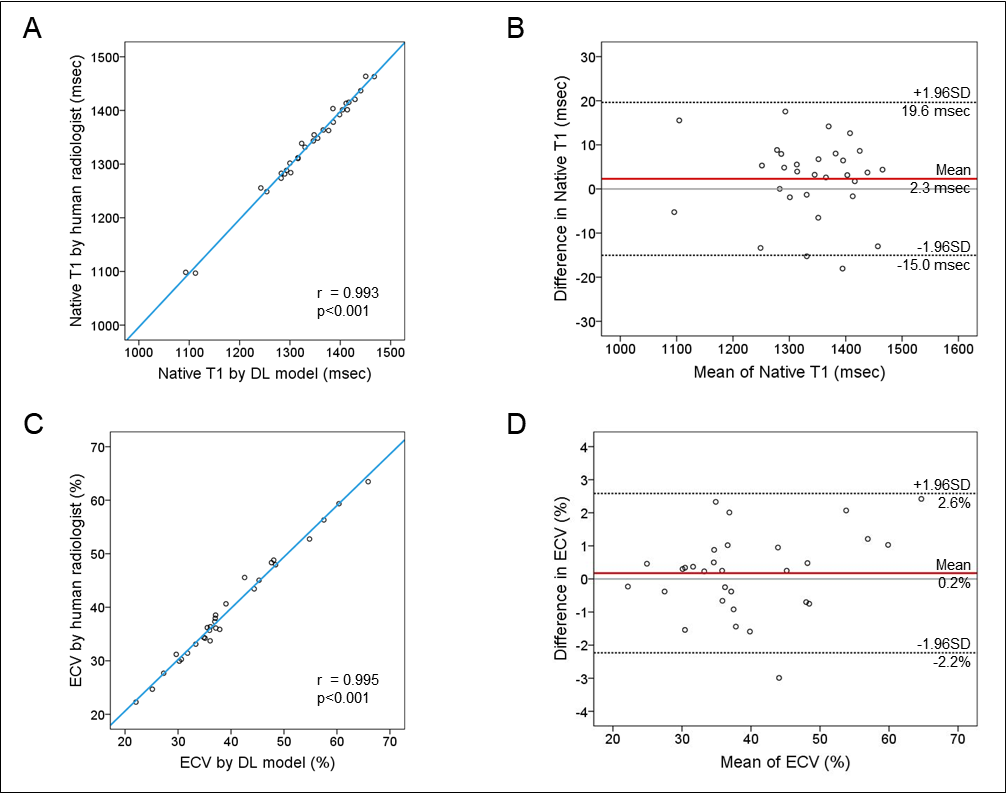

Correlations and agreements between the automated DL-based vs. manual (human radiologist) native T1 and ECV measurements are shown.

Abbreviations: DL, deep learning; SD, standard deviation; ECV, extracellular volume fraction

**Figure S3. ROC curves for the differentiation of CA from other etiologies of LVH using AI-derived native T1 and ECV**

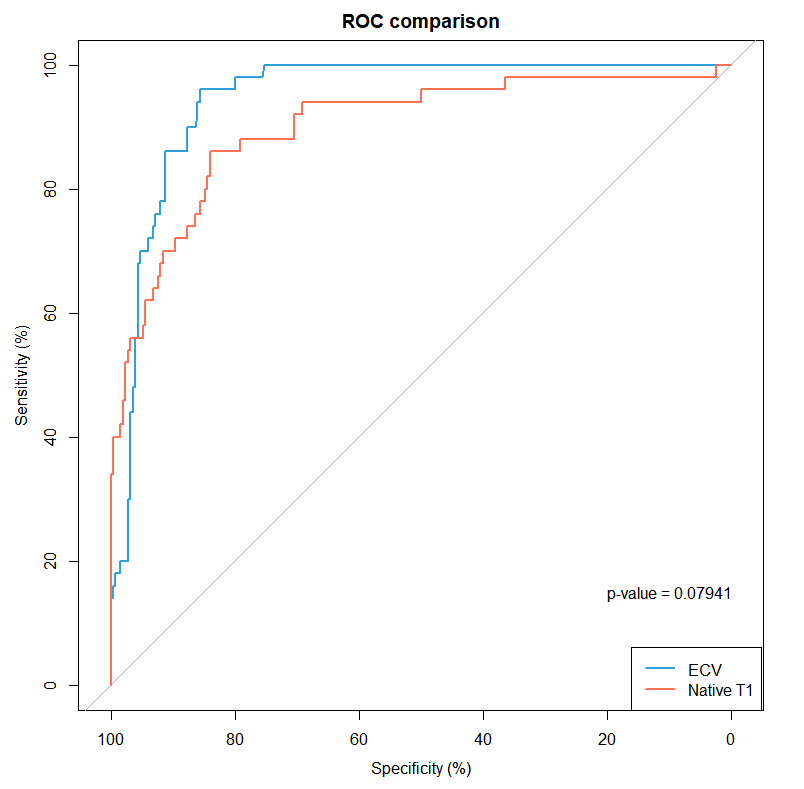

The diagnostic performance of the AI-derived native T1 and ECV measurements for the differentiation of CA from other etiologies of LVH is shown in the ROC curves.

Abbreviations: ROC, receiver operating characteristic; AI, artificial intelligence; ECV, extracellular volume fraction; AUC, area under the curve; CA, cardiac amyloidosis; LVH, left ventricular hypertrophy.

**Figure S4. ROC curves for the prediction of clinical outcomes of AL-CA using the AI-derived native T1 and ECV**

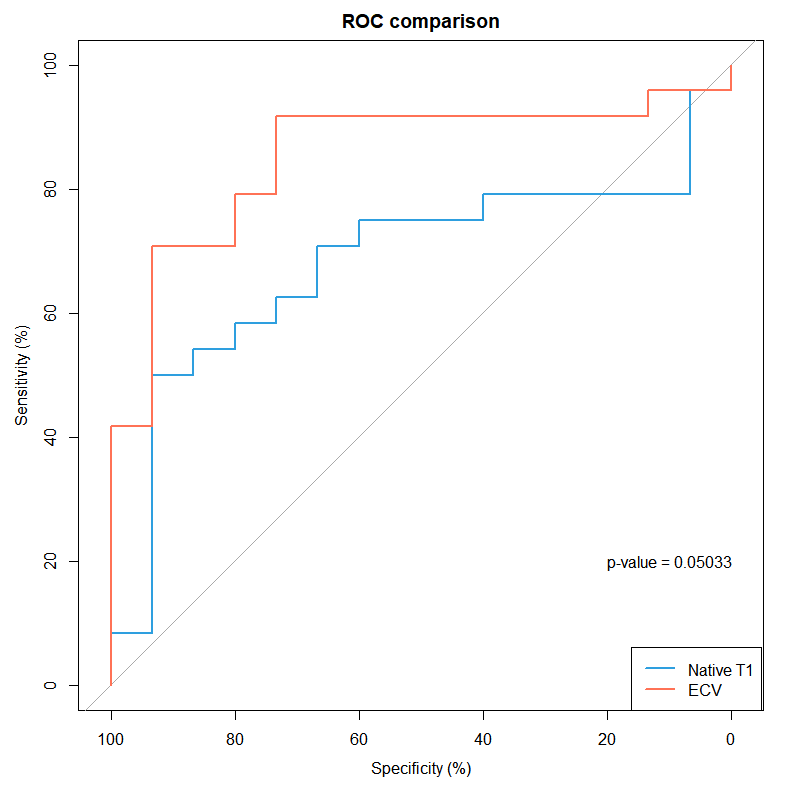

The performance of the AI-derived native T1 and ECV measurements in the prediction of cardiovascular death and hospitalization for heart failure among patients with AL-CA is shown in the ROC curves.

Abbreviations: ROC, receiver operating characteristic; AI, artificial intelligence; ECV, extracellular volume fraction; AUC, area under the curve; CA, cardiac amyloidosis; LVH, left ventricular hypertrophy.

**III. Supplementary Tables**

**Table S1. Univariable predictors of the composite outcome among patients with AL-CA**

|  | **Unadjusted HR (95% CI)** | **P-value** |
| --- | --- | --- |
| **Age (per +1 year)** | 0.953 (0.910 – 0.999) | 0.044 |
| **Age ≥60 years** | 0.100 (0.035 – 0.288) | <0.001 |
| **Age ≥65 years** | 0.529 (0.225 – 1.245) | 0.145 |
| **Age ≥70 years** | 0.768 (0.344 – 1.713) | 0.519 |
| **Male sex** | 1.012 (0.440 – 2.328) | 0.978 |
| **Hypertension** | 0.627 (0.249 – 1.583) | 0.323 |
| **Diabetes mellitus** | 0.702 (0.262 – 1.881) | 0.481 |
| **Chronic kidney disease** | 0.771 (0.319 – 1.863) | 0.564 |
| **Body-mass index (per +1 kg/m^2^)** | 0.914 (0.783 – 1.067) | 0.253 |
| **Systolic BP (per +1 mmHg)** | 0.967 (0.944 – 0.991) | 0.007 |
| **Systolic BP <100 mmHg** | 1.996 (0.844 – 4.720) | 0.116 |
| **Systolic BP <110 mmHg** | 3.114 (1.359 – 7.133) | 0.007 |
| **Diastolic BP (per +1 mmHg)** | 0.981 (0.949 – 1.015) | 0.266 |
| **Hemoglobin (per +1 g/dL)** | 1.075 (0.809 – 1.428) | 0.619 |
| **Hematocrit (per +1%)** | 1.016 (0.920 – 1.121) | 0.756 |
| **Glomerular filtration rate (per +1 mL/min/1.73m^2^)** | 1.007 (0.990 – 1.025) | 0.422 |
| **Albumin (per +1 g/dL)** | 0.661 (0.303 – 1.443) | 0.299 |
| **Delta FLC (per +1)** | 1.000 (0.999 – 1.001) | 0.615 |
| **Delta FLC ≥18 mg/dL** | 1.517 (0.566 – 4.066) | 0.408 |
| **NT-proBNP (per +1 pg/mL)** | 1.000 (1.000 – 1.000) | 0.073 |
| **NT-proBNP ≥1800 pg/mL** | 1.885 (0.442 – 8.046) | 0.392 |
| **CKMB (per +1 ng/mL)** | 0.996 (0.871 – 1.138) | 0.947 |
| **Troponin I (per +1 ng/mL)** | 0.999 (0.591 – 1.688) | 0.997 |
| **Troponin T (per +1 ng/mL)** | 235.334 (2.722 – 20348.545) | 0.016 |
| **Troponin I ≥0.1 ng/mL** | 1.593 (0.593 – 4.282) | 0.356 |
| **Troponin T ≥0.025 ng/mL** | 0.433 (0.146 – 1.286) | 0.132 |
| **Elevated troponin*** | N/A |  |
| **Native T1 (per +1 msec)** | 1.007 (1.001 – 1.012) | 0.021 |
| **Post T1 (per +1 msec)** | 0.994 (0.988 – 0.999) | 0.029 |
| **ECV (per +1%)** | 1.097 (1.045 – 1.152) | <0.001 |
| **T2 (per 1 msec)** | 1.041 (1.003 – 1.080) | 0.033 |
| **LV-EDV (per +1 mL)** | 1.005 (0.983 – 1.027) | 0.662 |
| **LV-EF (per +1%)** | 0.956 (0.912 – 1.002) | 0.061 |
| **LV-EF <50%** | 1.454 (0.538 – 3.931) | 0.461 |
| **LV-EF <55%** | 2.277 (1.012 – 5.122) | 0.047 |
| **LV-EF <60%** | 0.999 (0.437 – 2.285) | 0.998 |
| **LV-MI (per +1 g/m^2^)** | 1.005 (0.993 – 1.017) | 0.381 |
| **LAVI (per +1 mL/m^2^)** | 0.999 (0.973 – 1.026) | 0.946 |
| **E/e’ (per +1)** | 1.008 (0.966 – 1.052) | 0.722 |
| **TR Vmax (per +1 m/sec)** | 1.008 (0.967 – 1.051) | 0.703 |
| **LV-GLS (per +1%)** | 0.832 (0.724 – 0.955) | 0.009 |
| **LV-GLS <8%** | 2.484 (1.104 – 5.591) | 0.028 |
| **LV-GLS <10%** | 2.632 (1.039 – 6.666) | 0.041 |
| **LV-GLS <12%** | 3.269 (0.764 – 13.976) | 0.110 |
| **Revised Mayo stage (IV vs. III)** | 1.846 (0.764 – 4.465) | 0.173 |
| **ECV ≥40%** | 6.470 (1.910 – 21.914) | 0.003 |
| **ECV ≥45%** | 4.214 (1.789 – 9.926) | 0.001 |

* Elevated troponin was defined as either troponin T ≥0.025 ng/mL or troponin I ≥0.1 ng/mL.

Abbreviations: HR, hazard ratio; CI, confidence interval; BP, blood pressure; FLC, free light chain; NT-proBNP, N-terminal proB-type natriuretic peptide; CKMB, creatine kinase-myocardial band; ECV, extracellular volume fraction; LV, left ventricular; EDV, end-diastolic volume; EF, ejection fraction; MI, mass index; LAVI, left atrial volume index; TR, tricuspid regurgitation; GLS, global longitudinal strain; N/A, not applicable.
